# Direct Simulation of the CoVid-19 epidemic

**DOI:** 10.1101/2021.01.14.21249829

**Authors:** Augusto Cabrera-Becerril, Pedro Miramontes, Raúl Peralta

## Abstract

We introduce an agent-based model to simulate the epidemiological dynamics of COVID-19. Most computational models proposed to study this epidemic do no take into account human mobility. We present a direct simulation model where mobility plays a key role and propose as well four quarantine strategies. The results show that the no-quarantine strategy does lead to a high peak of contagions with no rebound. Quarantined strategies, for their part, show a re-emergence of the epidemic with smaller and softer peaks.

## 1. Introduction

The COVID-19 pandemic has affected more than twenty three million people in the world by August 25th, 2020. The first cases were reported in December 2019 in Wuhan, China. Currently (end of August, 2020) the infection per million inhabitants is relatively small (59 per million). However, other scenarios have been observed in different regions of the world. For example, in Mexico, there are around 4500 cases per million [13]. These data divergence can be explained by the containment measures applied for each country and even by the prevalence of co-morbidities associated with the infection. Such is the case of diabetes and hypertension [7]. In the lack of antiviral drugs and vaccines the only effective control policy seems to be the social distancing [2]. So far, data with the prevalence of covid-19 cases are widely available and allow to compare the evolution of the epidemic in different countries. In the present report, we propose a direct simulation model which takes into account human mobility and different quarantine policies as well as different intrahospital dynamics.

## 2. The Model

### 2.1. Background

Mathematical and computational modeling in epidemiology has a long tradition of mutual benefit between mathematics and medicine. Early attempts to use mathematical formalism to understand or to take actions against diseases date from the XVII century [5].

Since the seminal work of McKendrick and Kermack the so-called SIR models (“Susceptible Infectious Recovered) have been used with some degree of success. The class of SIR models are normally systems of ordinary deferential equations. As any ODE model, the underlying assumptions are, among many, a spatial homogeneous scenario The so called *well-mixed system*), a discrete population represented by continuous variables. Under special conditions SIR models have proven to be useful to predict some epidemic outbreaks [5].

There are many extensions of the SIR model in order to circumvent its restrictive underlying assumptions. Partial differential equations to take onto account spatial inhomogeneity [1], cellular automata to emphasize the discreetness of the individuals and the contact from person to person as the main contagion mechanism, dynamics over networks to include long range individuals transportation and some hybrid models [1].

Agent-based modeling (ABM) stands for a class of computational models that describes the dynamics of a many particles system using individual interactions that we implement to simulate the current epidemic.

### 2.2. The model

Our model, from now on CovABM, is an agent-based model for epidemic dynamics of covid-19 disease in urban centers or megalopolis. It aims to simulate transmission and disease dynamics including health care dynamics.

Agents represents people. In our model, in sharp contrast with SIR type models, human mobility plays an important role. The infection is by individual to individual contact but the individuals in the population are moving all the time.

In the first instance of our model the population, assumed to be constant, is *N*. In a first instance, there are 4*M* zones where these people inhabit. *M* is a parameter that may be chosen at will.

The state of each individual as a function of time is characterized as a vector of traits. Its state variables are age, comorbidities, the infection duration, if they are sick, if they show symptoms and their type of symptoms (mild, moderate, severe or critical).

There are also global properties of the system: the mean disease duration,the threshold of population for starting quarantine, the quarantine strategy, the capacity of health-care system (number beds for ICU patients).

At a certain time individuals are in one of the following four classes:

1. Susceptible (healthy)
2. Infected-asymptomatic
3. Infected-symptomatic
4. Cleared (deceased or recovered)

The dynamics of the model is based on local rules, each agent is the center of a Moore neighborhood where the infection dynamics actually occurs. A Moore neighborhood includes the agent itself, its neighbors at the four cardinal points and the four neighbors along the two diagonals. A healthy-susceptible person may be infected by an infected (symptomatic or asymptomatic) neighbor with a probability *P*. If the infected agent shows symptoms it may be at one of four stages of disease: mild (the majority of cases are of this degree and their symptoms are fever, cough, and shortness of breath. These symptoms appear between 0 and 14 days after infection. These patients do not require hospitalization or specific treatment), moderate (same symptoms as mild, but are persistent, usually up to seven days and worsen over time. Some therapeutic options can be effective in these cases), severe (to the above symptoms are added mental confusion and bruising in the extremities and lips. In these cases hospitalization is necessary) and critical (In these cases patients cannot breathe. Pneumonia symptoms are obvious and an intensive care unit is required).

Mild symptomatic agents have two possible outcomes: either to clear-out or to evolve to moderate. Moderate also have two possibilities: to clear-out or evolve to a severe symptomatology. Severe cases may also clear-out or become critical. Critical cases either clear-out or die.

The infection dynamics depends on the mobility of individuals. Agents move following levy-flights among nearby urban zones, the length of each flight is distributed among the populations in such manner that several individuals are constrained to local mobility and few of them have long trajectories.

Thus the Moore neighborhood of each agent changes dynamically over the time. Asymptomatic infected individuals that have longest trajectories are more likely to infect more people.

The quarantine is modeled as an external stimulus, modeled by setting a threshold for infected symptomatic population (the *qt* parameter), there are three distinct strategies for quarantine:

1. Strategy 1 consists on restricting mobility only of symptomatic infected.
2. Strategy 2 consist on restricting mobility of approximate 30 % of the population.
3. Strategy 3 consist on restricting mobility of approximate 80 % of the population.

We include a responsiveness parameter that measures how disciplined are people in following the instructions of the health authorities. Also the efficacy of information given by the Health authorities is measured by an index. Both information-efficacy and responsiveness are numbers between 0 and 1. Responsiveness distributes over the population and we variate information-efficacy on the numerical experiments.

We ran several numeric experiments, with four distinct strategies: No-quarantine, and quarantine strategies 1-3. We variate the parameter **qt** that measures the quarantine threshold with values 15, 5, and 50. The parameter **mb** that measures the maximum amount of beds per hospital with values 50, 70 and 90. The information efficacy take values on 0.3, 0.5 and 0.8. We consider 16 urban centers with total population of 8000 individuals. We did a 365 days cycle in each run. And 3200 runs of each experiment.

Severe and critical symptomatic individuals are hospitalized as long as the health care capacity is not exceeded. In such a case, there are no more beds in hospitals and the individuals in severe or critical conditions are more likely to die.

## 3. Results

The numerical experiments in figure 4 show that in the non-quarantine strategy the number of infected decreases, while those recovered increase over time. However, in quarantine strategies, the curves of recovered and infected never cross, that is, the infected do not increase in number over time. Figure 5 shows the comparisons between the different quarantine and non-quarantine strategies over time. Clearly, the non-quarantine strategy maintains a high number of infected over time, while quarantine measures decrease the number of infected over time. The most important qualitative change is shown in figure 6. In the non-quarantine strategy, the epidemic does not re-emerge over time, but maintains a high number of infected, while the quarantine strategies re-emerge over time, but keeping a low number of infected.

**Figure 1.**
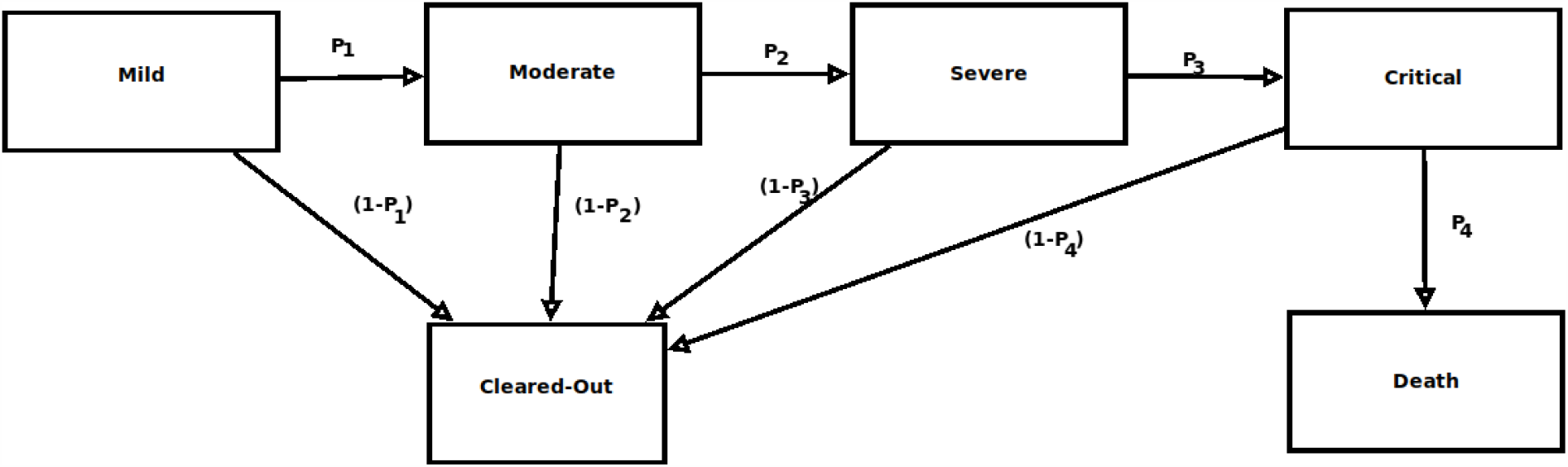
Dynamics for the CoVid-19 Disease. The diagram depicts the overall evolution of the disease from mild cases to several and critical ones. The *P*_*i*_ stands for the probability of state transition, in each case, (1−*P*_*i*_) stands for the probability for each individual to clear-out given its previous state of disease.

**Figure 2.**
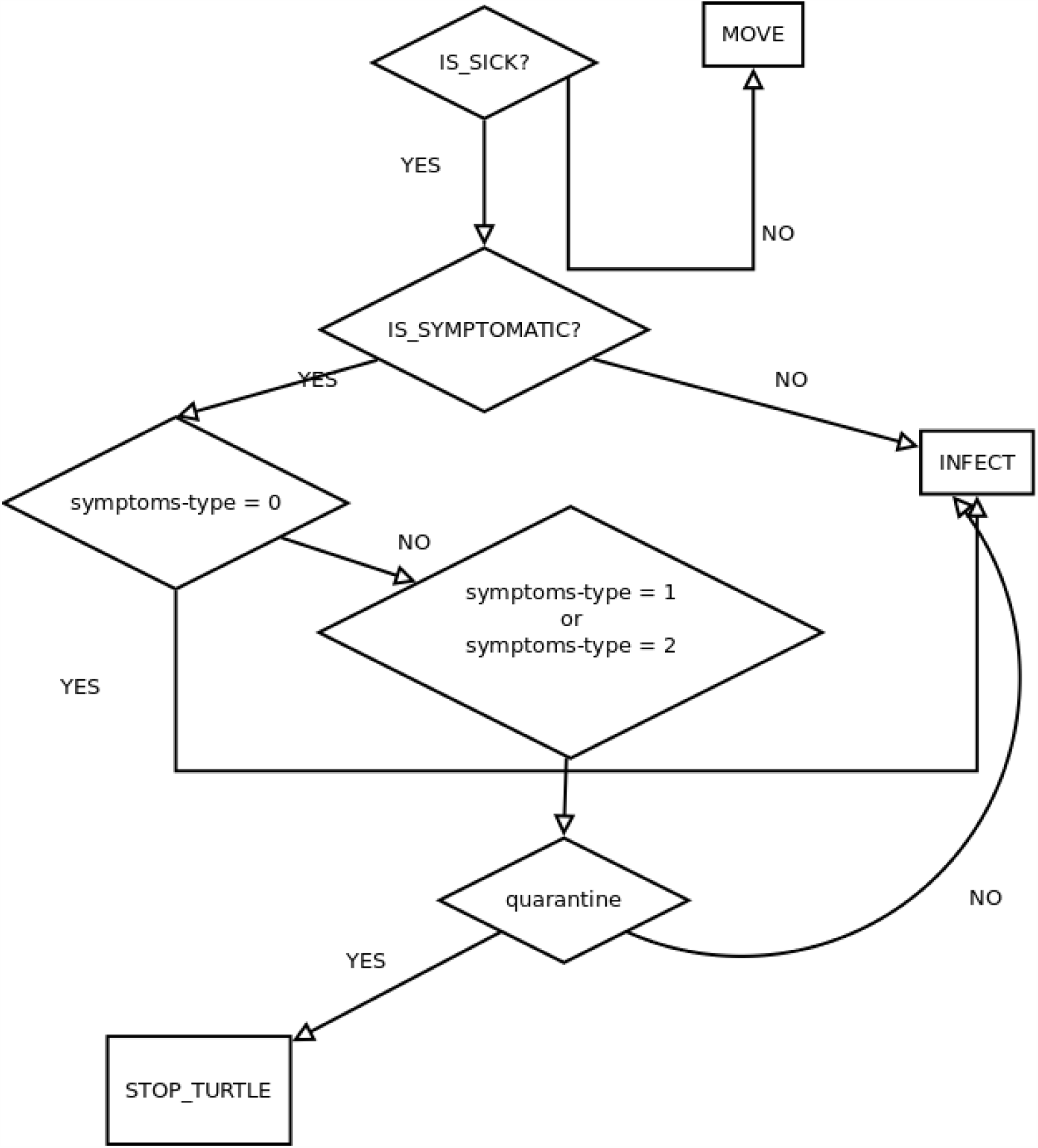
Infection dynamics flowchart. This diagram explains the steps that an infected agent takes to infect other agents in its neighborhood.

**Figure 3.**
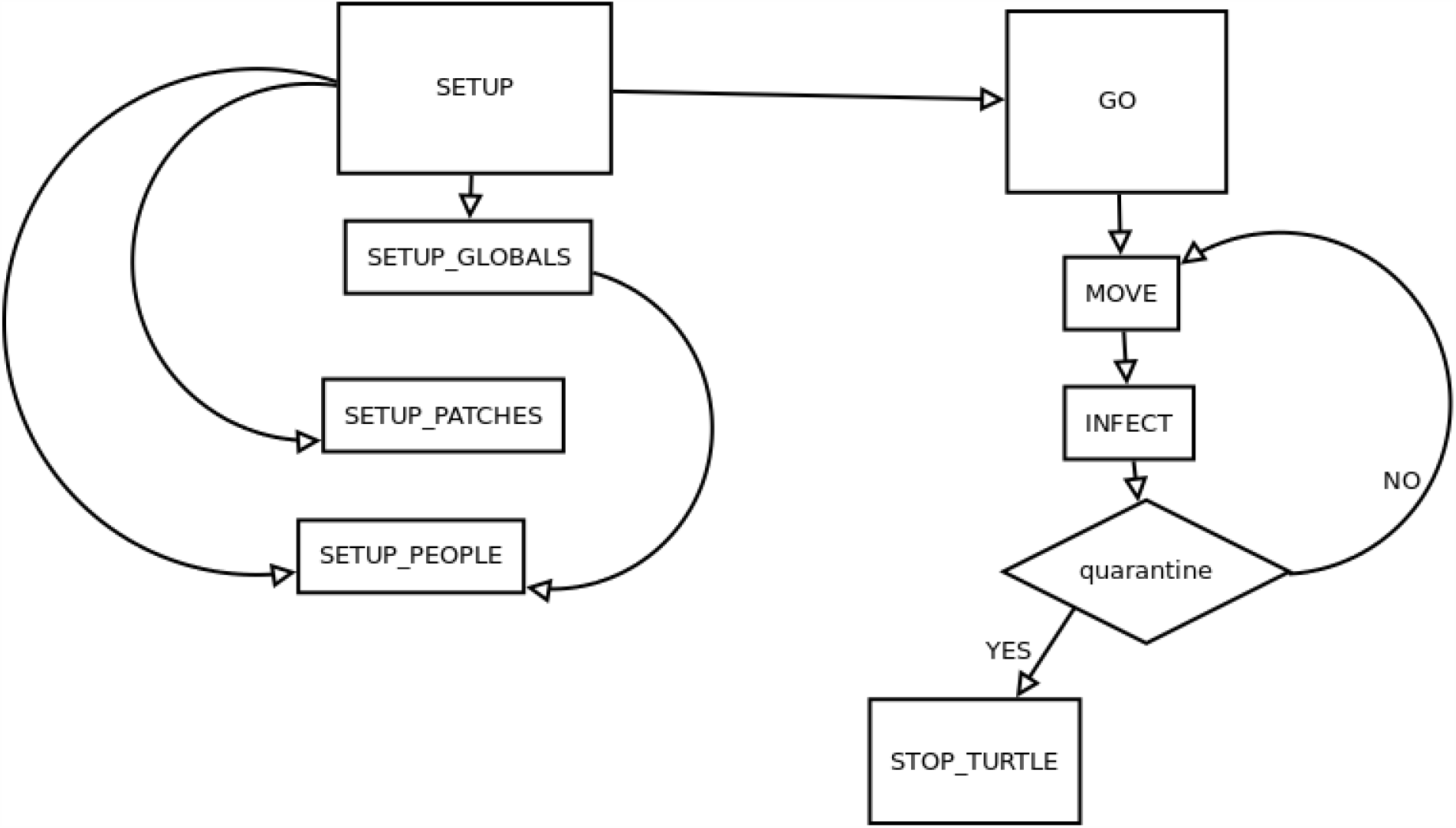
CovABM model Flow chart. Our model has two main methods. “SETUP” initializes the global, patches and agent variables. “GO” contains the agent methods on how the agent moves, how infect their neighbors.

**Figure 4.**
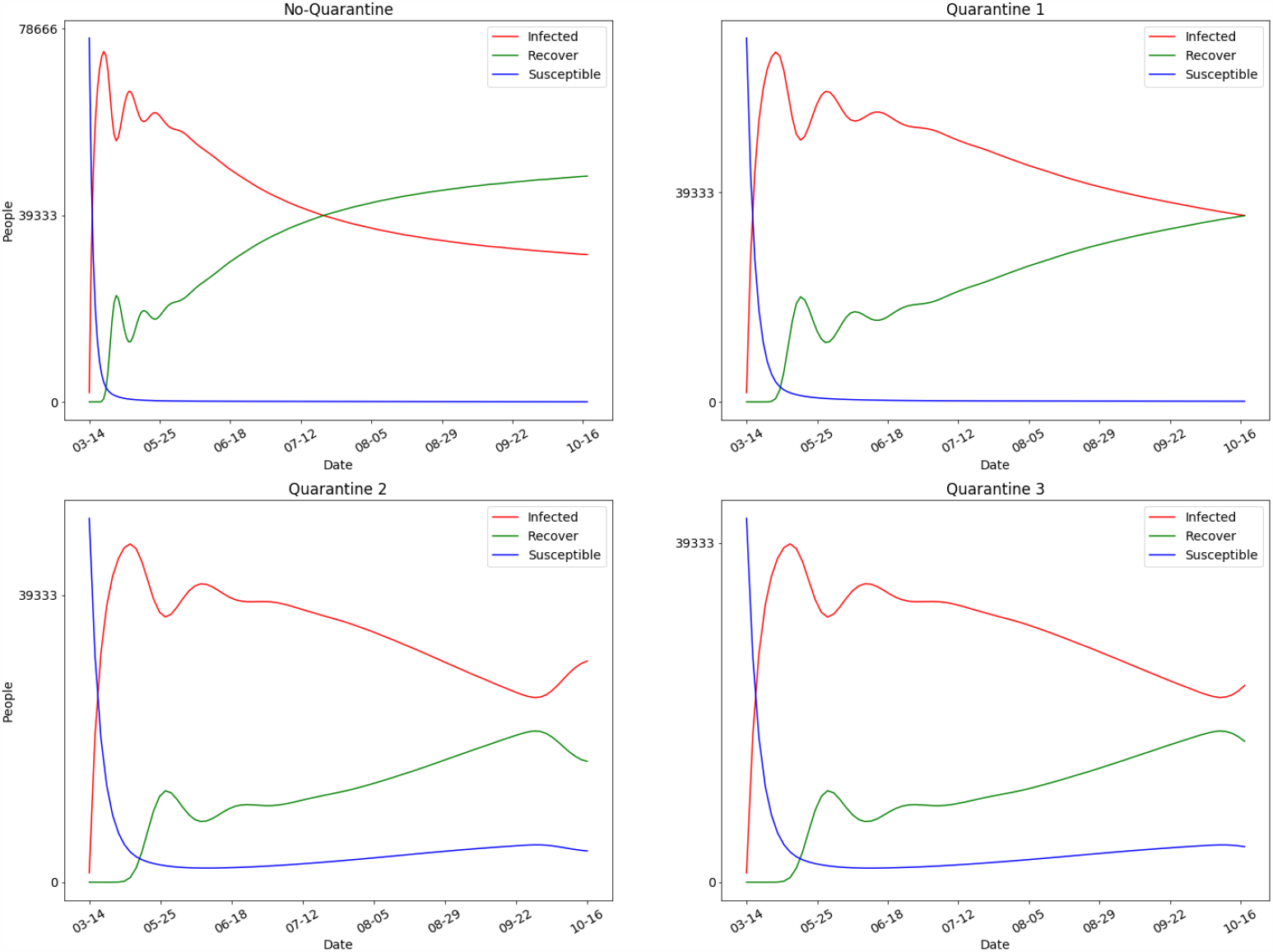
Mean values from 2400 runs of numerical experiments. In the plots above we show mean values for susceptible, infected and recovered individuals in each of four quarantine scenarios. Population is represented on the x-axis and time in weeks on the y-axis. The most evident qualitative difference is shown between the first scenario (not quarantine) where the infected and recovered curves intersect and the following scenarios (quarantine) where the infected and recovered curves do not intersect.

**Figure 5.**
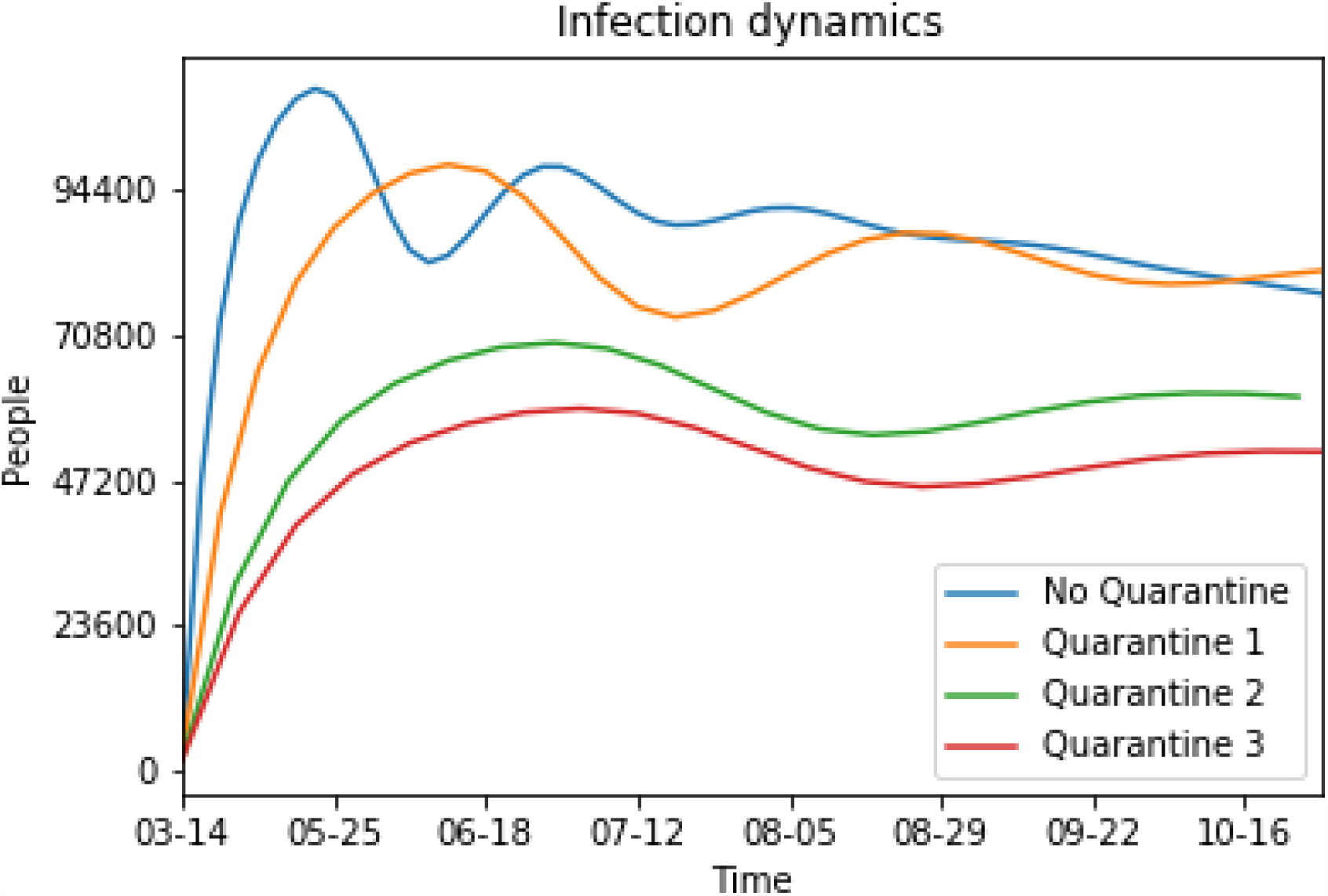
Comparison between different epidemic scenarios. In this plot we show mean values for infected individuals, comparing four different quarantine scenarios. Population is represented on the x-axis and time in weeks on the y-axis. Strategy 1 and No-Quarantine cases are similar, although in strategy 1 the maximum of infected is below the No-quarantine as expected, in about 4 months both curves are very similar. Strategies 2 and 3 are very similar overall time. This figure shows the case where strategies 2 and 3 clearly maintain a lower number of infected over time.

**Figure 6.**
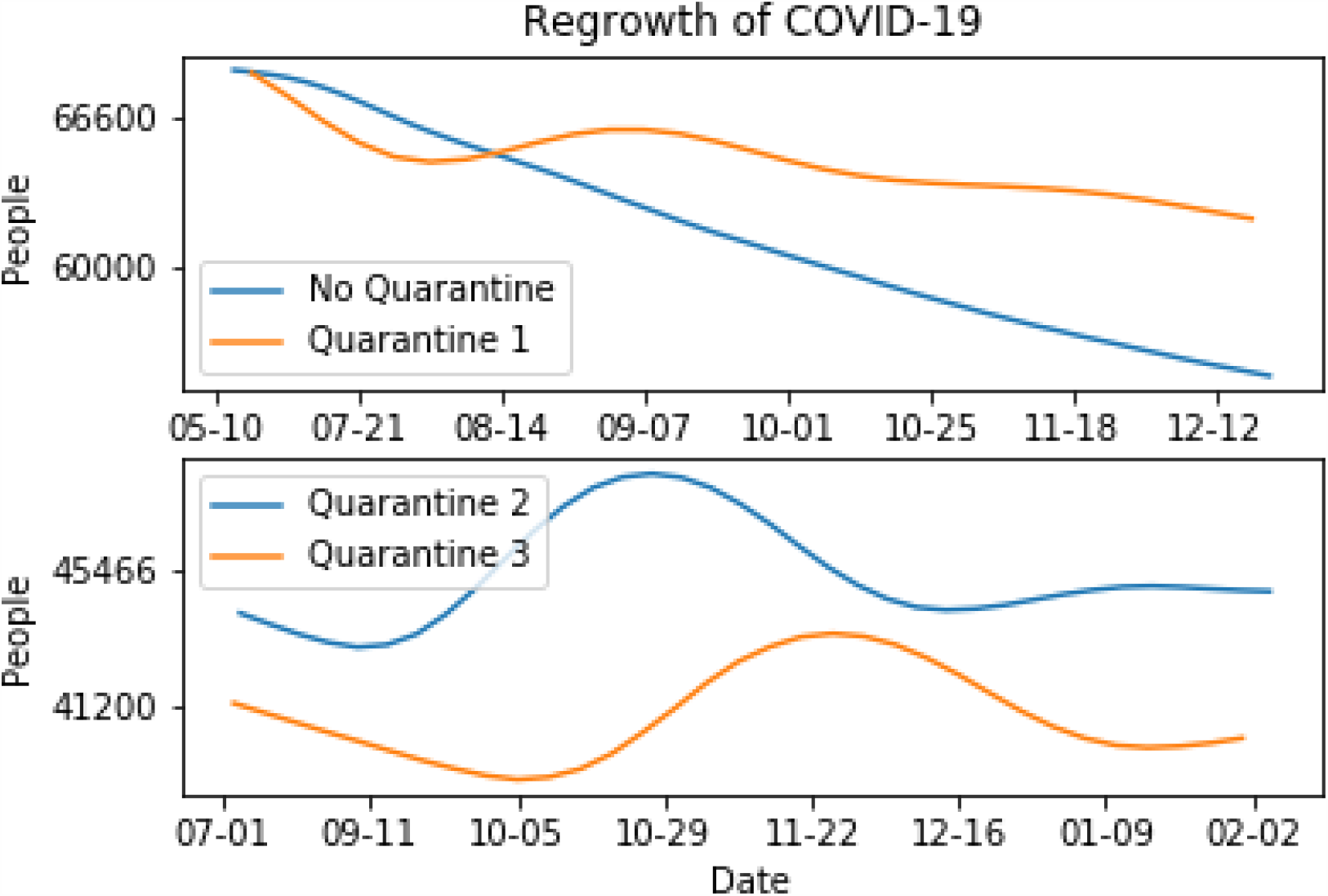
Regrowth in four different scenarios. In these plots we compare re-growth for No-quarantine strategy and strategy 1 also for quarantine strategy 2 and strategy 3. Strategies 2 and 3 seem more effective that strategy 1. Strategy 1 shows a more abrupt regrowth than other two strategies or even the No-quarantine case.

## 4. Discussion

Mathematical modeling has been widely used in the current COVID-19 pandemic. The most employed scheme in the present circumstances is the SIR models and its variants with the subsequent calculation of the basic reproductive number (R0). Despite the efforts of many teams around the world, these family of models have failed to produce sound results [9]. The assumptions behind SIR models are too strong to allow trustworthy predictions. The hardest constraint is to assume an spatial homogeneous population distribution. What is something hard to accept. Several efforts have been made, with different levels of success, using data-driven models based on neural computation [3]. It is our belief that no model of epidemic would be reliable if it does not take into account human mobility.

On the other hand, direct computational simulation is a powerful, brute force, tool that circumvents that problem. On the other hand, direct simulation models require a huge amount of computing. Perhaps it would be a good collective effort to join forces leading to the organization of groups using the tools of distributed computing to tackle this problem in a collaborative way. Since the detection of this new disease in humans, multiple efforts have been made to understand the biology of the causative agent (SARS-CoV-2), as well as its human-to-human transmission mechanisms. In this effort, the virus receptor (ACE2) could be determined. Such finding has allowed designing therapeutic strategies aimed at blocking this receptor. Likewise, the study of the genomic structure of the virus and its replication mechanism mediated by the enzyme replicase, has allowed the design of therapeutic strategies to inhibit replicase. In this way, the study of the biology of the virus has allowed the design of specific therapeutic strategies [10]. However, because these strategies are not yet available, the best way to control the spread of the disease has been through social isolation or quarantine strategies. Practically all the countries affected by the pandemic, established the quarantine strategy, to avoid saturation of their health systems. In our model we consider three quarantine strategies and one non-quarantine strategy. Our results show that the non-quarantine strategy has a dynamic with a single very large epidemic outbreak that would have saturated the health system. Although some authors maintain that this strategy could have contributed to overcoming the pandemic due to herd immunity [4]. On the other hand, quarantine strategies show a smaller epidemic outbreak, both in the mobility restriction strategy only for symptomatic people, and in the mobility restriction strategy for 30 and 80 of the population. These strategies may not saturate health systems, but they depend on people restricting their mobility. Interestingly, these strategies show a re-emergence of the epidemic when restriction measures are relaxed. We do not discard that this new outbreak would be minor or null with the introduction of specific therapies.

Our model does not pretend to be accurate in the quantitative predictions but the outcome is pretty consistent qualitatively with what the data shows by the end of August. As far as we know, there is no a single country which has not applied no quarantine at all. Examples of countries with a very loose quarantine (orange line in our Figures 5)are the United States, UK and Sweden. Even if they corrected their strategy later, the outcome was somehow determined by the first stages of their local epidemic. Mexico and Germany are examples of the quarantine-3 (green line). They applied a rather strict quarantine with a later gradual release and subsequent opening of the economy. Finally, the red line in Figure, a severe and strict population confinement, is illustrated by China and Australia.

## Data Availability

The authors confirm that the data supporting the findings of this study are available within the article [and/or] its supplementary materials.

## Acknowledgment

This study was partially supported by DGAPA-PAPIT grant IN-345512. The authors thank the High Performance Computing Lab, Faculty of Sciences, UNAM.

---

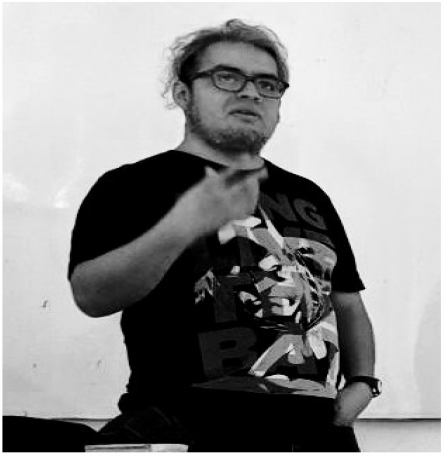

**Augusto Cabrera-Becerril** - graduated from the Faculty of Sciences at UNAM, Mexico.He obtained a MsSc at the same university working on Agent-Based Modeling under the advise of Pedro Miramontes. Currently he is a lecturer at the Faculty of Sciences at UNAM

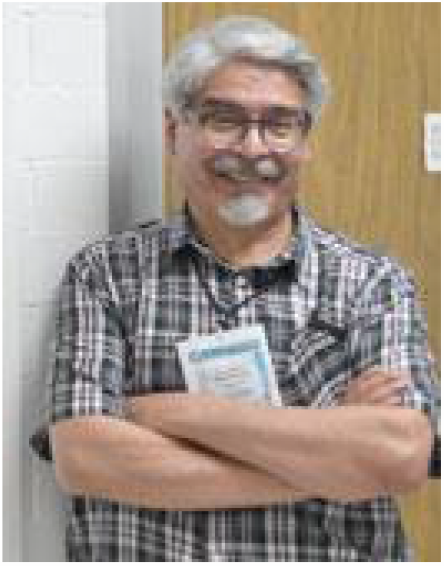

**Pedro Miramontes** - studied physics at the Faculty of Sciences, UNAM. He received a PhD degree in Mathematics from the same university. Since 1975 he teaches at the same Faculty.

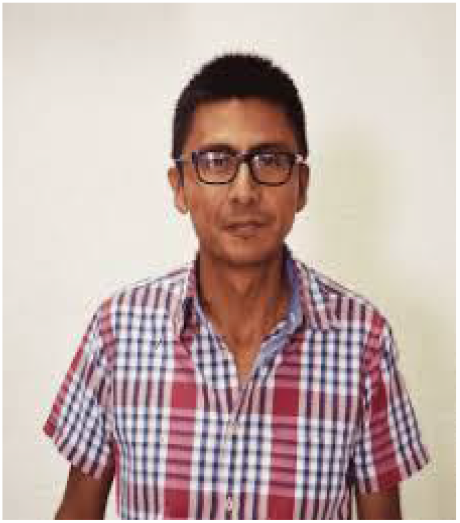

**Raúl Peralta** - graduated from the Faculty of Medicine at UNAM, Mexico. He obtained a PhD at the same university working in a viral infection control at the Social Security Mexican Institute. Currently he is professor at the Autonomous University of the State of Morelos

